# Public oral health screening leads to regular dental visits: the LIFE Study

**DOI:** 10.1101/2023.09.07.23295186

**Authors:** Yudai Tamada, Taro Kusama, Megumi Maeda, Fumiko Murata, Ken Osaka, Haruhisa Fukuda, Kenji Takeuchi

**Affiliations:** Department of International and Community Oral Health, Tohoku University Graduate School of Dentistry, Miyagi, Japan; Department of Preventive Medicine, Nagoya University Graduate School of Medicine, Aichi, Japan; Division of Statistics and Data Science, Liaison Center for Innovative Dentistry, Tohoku University Graduate School of Dentistry, Miyagi, Japan; Department of Health Care Administration and Management, Kyushu University Graduate School of Medical Sciences, Fukuoka, Japan

**Keywords:** Dental check-up, Dental screening, Individual screening, Dental care utilization

## Abstract

**Background:** Although periodontal disease screening has been implemented as a public oral health screening program in Japan, it remains unclear whether screening encourages regular dental visits. This study aimed to test the hypothesis that periodontal disease screening leads to subsequent regular dental visits among adults using a difference-in-differences (DID) approach.

**Methods:** This study used healthcare claims data of municipality residents who underwent periodontal disease screening in 2017 or 2018. For each screening recipient, four individuals of the same age and sex were extracted from those who did not undergo screening as controls. In the DID analysis, we assessed the change in the prevalence of dentist visits at least once every 180 days after screening.

**Results:** A total of 4,270 participants were included in the analysis. The prevalence of visiting dentists was consistent before screening among the participants who underwent screening (181–360 days before, 62.4%; 1–180 days before, 60.3%). While the prevalence was also consistent in those who did not undergo screening throughout the study period (approximately 45%), the prevalence in those who underwent screening sharply increased after undergoing the screening (1–180 days after, 81.1%) and then declined (181–360 days after, 67.8%). DID analysis indicated that the prevalence increased by 12.1% owing to screening. In addition, the age-subgroup DID estimates were higher in the younger population (aged 20–35 years, 17.4%; 40–55 years, 11.5%; 60 years, 11.0%).

**Conclusion:** Periodontal disease screening increased subsequent dental visits, especially in younger populations.

## INTRODUCTION

Oral diseases are among the most prevalent health conditions that impose tremendous health and economic burdens^1^. Periodontal disease, a representative oral disease, is a known risk factor for various systemic diseases, including dementia^2^, cardiovascular diseases^3^, and pulmonary diseases^4,5^, through sharing a common inflammatory pathway^6^. Regular dental visits contribute to the early detection of oral diseases, and continuous preventive care by dental professionals is effective in maintaining and improving oral health. Previous studies reported that preventive dental visits reduce future non-preventive dental care^7^ and dental expenditures^7,8^. This suggests that health policy should prioritize frequent dental visits to improve people’s health.

Various public oral health screening programs have been implemented in Japan. Periodontal disease screening is one of them, and it is primarily aimed at middle-aged and young older populations, where at least 40% of dentate adults aged ≥40 years experience periodontal disease^9^. Screening was initially introduced in 1995 and is now conducted for adults aged 40, 50, 60, and 70 years as part of a health promotion project. Eligible people can undergo screening free of charge or at a low cost (depending on their residential municipality)^10^. During screening, people will be examined for their oral status, such as the condition of present teeth, tooth loss, and periodontal tissue, by the procedure set out^11^. Based on the examined results, they will receive oral hygiene education, such as the importance of regular dental visits and advice on improving their oral status. Many companies have private workplace oral health promotion aimed at a similar age group as those included in the periodontal disease screening program. Such programs, which consist of oral examinations and tailored oral health instructions, have previously been reported to increase the frequency of dental visits^12,13^. Although the purpose and content of periodontal diseases are similar, it is still unknown whether periodontal disease screening can also contribute to an increase in subsequent dental visits. Therefore, this study aimed to test the hypothesis that screening for periodontal disease leads to subsequent regular dental visits among adults. Using the difference-in-differences (DID) approach, we assessed changes in the prevalence of regular dental visits after periodontal disease screening.

## METHODS

### Study Population and Setting

This study was based on data from the Longevity Improvement & Fair Evidence (LIFE) Study^14^, a longitudinal community-based database project that collected various health-related data, such as administrative healthcare claims and oral health screenings data, from municipal governments. In the LIFE Study, a unique research ID was assigned to each resident by data managers, and each data can be linked at the individual level. Details of the LIFE Study, such as the data collection procedures and database construction, are available^14^. In this study, we used data on periodontal disease screening conducted in fiscal years 2017 or 2018 and healthcare claims of the National Health Insurance^15,16^ enrollees collected from a municipality with a residential population of approximately 700,000.

In the municipality, periodontal disease screening is conducted for adults aged 20, 25, 30, 35, 40, 45, 50, 55, 60, 65, and 70 years from June to November in designated dental clinics as individual screenings free of charge. The municipality also had other oral health screening programs (i.e., oral healthcare screening), annually conducted for those aged ≥65 years throughout the year. Because those aged 65 or 70 years could undergo oral healthcare and periodontal disease screening within a year, we excluded them from our analytical sample to assess the effect of only periodontal disease screening. Additionally, those who underwent periodontal disease screening more than twice a year or those aged out of eligibility for the screening were excluded from the analysis. Furthermore, we restricted our analytic sample to those who had continuous health insurance enrollment for the 360 days preceding and following periodontal disease screening. We assessed the enrollment in the following two ways: First, in our primary analysis, eligible participants who visited a medical institution were regarded as having health insurance enrollment for 180 days since the visit date (when ≥181 days had passed since the last visit date, they were regarded as being disenrolled from health insurance at 180 days since the previous visit date). Second, in our secondary analysis, although the data existed only for those aged ≥40 years, we ascertained the participants’ health insurance enrollment period using the health insurance register collected from the municipality.

### Outcome Assessment

The primary outcome was the prevalence of participants who visited dentists at least once every 180 days for 360 days before or after undergoing periodontal disease screening (181–360 days before; 1–180 days before; 1–180 days after; and 181–360 days after undergoing screening). Dental visits were identified based on dental claims. The interval between regular dental visits (180 days) was determined because general dental practitioners traditionally recommend visiting a dental clinic at least once every 180 days to maintain oral health^17^. In the sensitivity analysis, to account for other treatment or maintenance strategies^18^, we assessed the prevalence of visiting dentists at least once every 90 days before or after undergoing screening (271–360 days before; 181–270 days before; 91–180 days before; 1–90 days before; 1–90 days after; 91– 180 days after; 181–270 days after; 271–360 days after undergoing screening).

Furthermore, to investigate the details of dental visits after undergoing the screening, for example, periodontal maintenance and caries treatment, we assessed the prevalence of the following seven dental procedures at least once in 1–180 or 181–360 days after undergoing the screening: cavity fillings, pulpectomies, dental calculus removals, periodontal surgeries, tooth extractions, dental bridges, and dentures. The procedures were identified using the original Japanese procedure codes in the dental claims data according to the definitions in previous studies^19,20^ (Supplemental Table 1).

### Exposure Assessment

The exposure in this study involved screening for periodontal disease. For each participant who underwent screening, four participants of the same age and sex were sequentially selected from those who did not undergo screening (1:4 age and sex matching was conducted) in a non-replicative manner.

### Statistical Analysis

First, for demographic characteristics, we assessed the participants’ age, sex, and screening month by fiscal year when they underwent periodontal disease screening. Second, we evaluated descriptively whether the prevalence of visiting dentists at least once in 180 days increased after periodontal disease screening (for participants who did not undergo screening, the matched date was regarded as the index date). We then examined the changes in the prevalence of visiting dentists 180 days before and after undergoing screening using the DID approach^21,22^ after statistically verifying the parallel trend assumption. The DID analysis allowed for a counterfactual assessment of what happened to the intervention group (those who underwent screening) after the intervention (undergoing screening) by referring to the time course of the control group (those who did not undergo screening). Practically, we included an interaction term between the indicator of whether the participants experienced the screening and the period after undergoing the screening in a linear probability model, and the coefficient was the DID estimate, which can be interpreted as the percentage change due to undergoing the screening. The 95% confidence intervals (CIs) for the DID estimates were obtained using wild cluster bootstrapping with 1,000 replications. As an additional analysis, we descriptively assessed whether the prevalence of visiting dentists within 90 days of screening increased. Furthermore, as an age-subgroup analysis, we evaluated the transitions in the prevalence of visiting dentists by age group (20–35/40–55/60 years). Third, we restricted the analytic sample to those who had not visited dentists 360 days before the screening and assessed the transition of the prevalence of visiting dentists in 180 days in the total population and the age groups (aged 20– 35/40–55/60 years). Fourth, we assessed the participants’ dental procedures at least once in 1– 180 days or 181–360 days after screening.

As a sensitivity analysis, we conducted the same analysis as the main analysis among participants whose continuous health insurance enrollment was ascertained from the health insurance register. All analyses were conducted using Stata (version 17.0; Stata Corp., College Station, TX, USA). This study followed the Strengthening the Reporting of Observational Studies in Epidemiology (STROBE) guidelines.

### Ethical Considerations

This study was approved by the Kyushu University Institutional Review Board for Clinical Research (approval number: 22114-02) and the Ethics Committee of Tohoku University Graduate School of Dentistry (approval number: 23835). Data usage approval was obtained from the municipality’s Personal Information Protection Review Board.

## RESULTS

Table 1 shows the characteristics of the analytical sample according to the fiscal year in which the participants underwent periodontal disease screening. Of 854 participants who underwent screening (427 in the fiscal year 2017 and 427 in the fiscal year 2018), 3,416 participants were matched with those who did not undergo screening. Among the participants, 64.8% were female, 11.6% were 20–35, 64.4% were 40–55, and 24.0% were 60 years old. In addition, the participants underwent screening most frequently in November (21.2%), followed by June (21.0%).

**Table 1.** Characteristics of analytic sample according to fiscal year when participants underwent periodontal disease screening.

|  | Overall<br>(n = 4,270) |  | Fiscal year 2017 |  |  |  | Fiscal year 2018 |  |  |  |
| --- | --- | --- | --- | --- | --- | --- | --- | --- | --- | --- |
|  |  |  | Participants who underwent periodontal disease screening<br>(n = 427) |  | Participants who did not undergo periodontal disease screening<br>(n = 1,708) |  | Participants who underwent periodontal disease screening<br>(n = 427) |  | Participants who did not undergo periodontal disease screening<br>(n = 1,708) |  |
|  | n | % | n | % | n | % | n | % | n | % |
| Sex |  |  |  |  |  |  |  |  |  |  |
| Male | 1,505 | 35.3 | 146 | 34.2 | 584 | 34.2 | 155 | 36.3 | 620 | 36.3 |
| Female | 2,765 | 64.8 | 281 | 65.8 | 1,124 | 65.8 | 272 | 63.7 | 1,088 | 63.7 |
| Age, years |  |  |  |  |  |  |  |  |  |  |
| 20 | 65 | 1.5 | 7 | 1.6 | 28 | 1.6 | 6 | 1.4 | 24 | 1.4 |
| 25 | 60 | 1.4 | 2 | 0.5 | 8 | 0.5 | 10 | 2.3 | 40 | 2.3 |
| 30 | 120 | 2.8 | 11 | 2.6 | 44 | 2.6 | 13 | 3.0 | 52 | 3.0 |
| 35 | 250 | 5.9 | 20 | 4.7 | 80 | 4.7 | 30 | 7.0 | 120 | 7.0 |
| 40 | 360 | 8.4 | 43 | 10.1 | 172 | 10.1 | 29 | 6.8 | 116 | 6.8 |
| 45 | 490 | 11.5 | 45 | 10.5 | 180 | 10.5 | 53 | 12.4 | 212 | 12.4 |
| 50 | 1,115 | 26.1 | 121 | 28.3 | 484 | 28.3 | 102 | 23.9 | 408 | 23.9 |
| 55 | 785 | 18.4 | 79 | 18.5 | 316 | 18.5 | 78 | 18.3 | 312 | 18.3 |
| 60 | 1,025 | 24.0 | 99 | 23.2 | 396 | 23.2 | 106 | 24.8 | 424 | 24.8 |
| Screening month |  |  |  |  |  |  |  |  |  |  |
| June | 179 | 21.0 | 84 | 19.7 | — | — | 95 | 22.3 | — | — |
| July | 164 | 19.2 | 82 | 19.2 | — | — | 82 | 19.2 | — | — |
| August | 118 | 13.8 | 60 | 14.1 | — | — | 58 | 13.6 | — | — |
| September | 101 | 11.8 | 54 | 12.7 | — | — | 47 | 11.0 | — | — |
| October | 111 | 13.0 | 52 | 12.2 | — | — | 59 | 13.8 | — | — |
| November | 181 | 21.2 | 95 | 22.3 | — | — | 86 | 20.1 | — | — |

Notes: In this analytical sample, participants' continuous health insurance enrollment was ascertained based on their healthcare utilization patterns (those who visited a medical institution were regarded as having health insurance enrollment for 180 days since the visit date).

Figure 1 presents the prevalence of visiting dentists at least once in 180 and 360 days before and after undergoing screening. In the total population, the prevalence before screening was almost consistent throughout the 360 days in both participants who underwent screening (181–360 days before, 62.4%; 1–180 days before, 60.3%) and those who did not (181–360 days before, 43.5%; 1–180 days before, 43.5%) (parallel trend test, *P* = 0.305). The prevalence was also almost consistent after undergoing screening in those who did not (1–180 days after, 44.1%; 181–360 days after, 45.0%). In contrast, in those who underwent screening, the prevalence sharply increased after screening (1–180 days after, 81.1%) and then declined (181– 360 days after, 67.8%) but remained higher than before screening. The DID analysis indicated that the prevalence was increased by 12.1% (95% CI, 9.0–15.0) by undergoing the screening (Table 2). In an additional analysis that assessed the prevalence of visiting dentists at least once in 90 days during the 360 days before and after undergoing the screening, a similar trend was observed: the prevalence sharply increased after the screening and then declined (Supplemental Figure 1).

**Figure 1.**
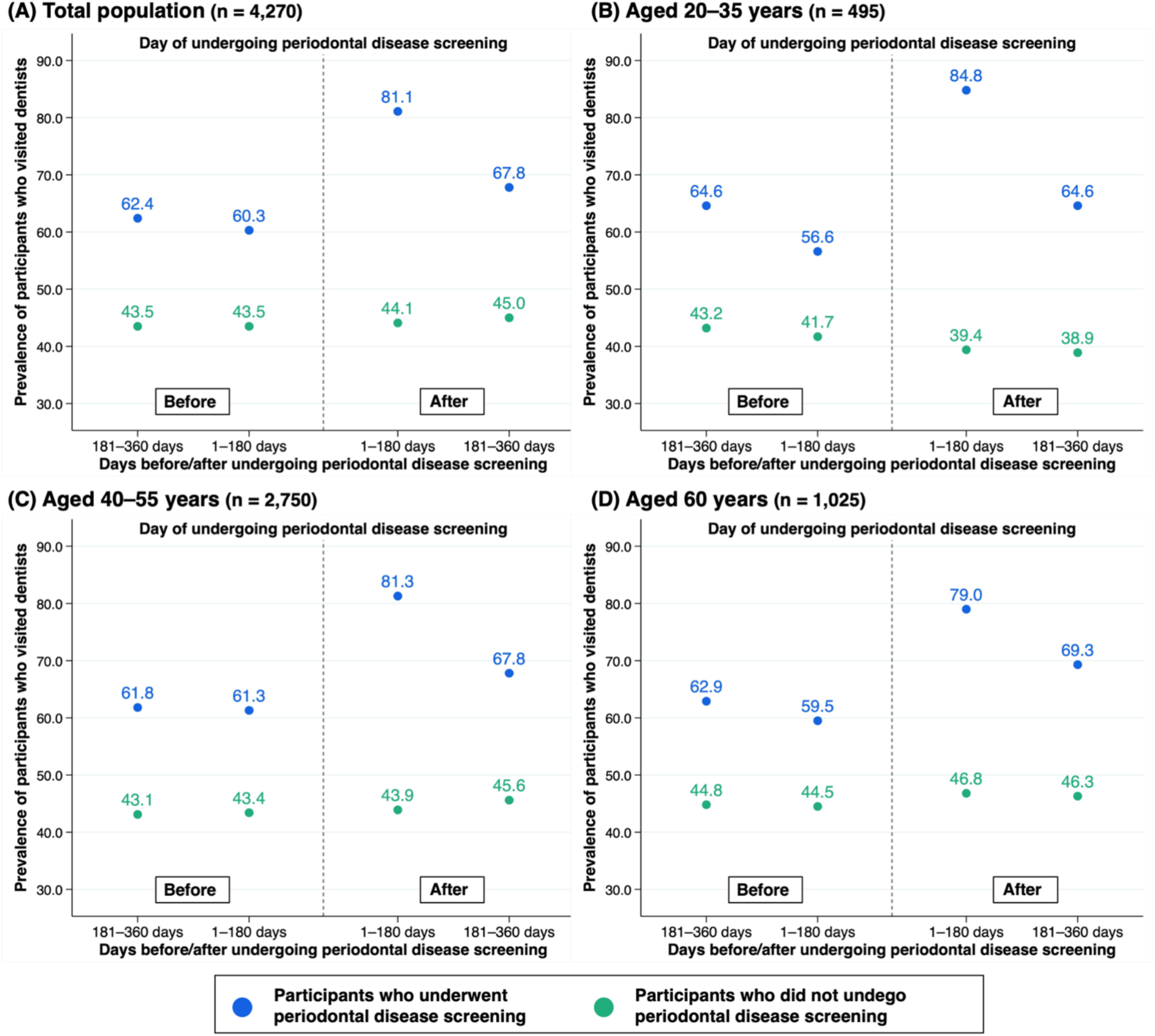
Prevalence of participants who visited dentists in 180 days during 360 days before and after undergoing periodontal disease screening in (A) total population, (B) aged 20–35 years, (C) 40– 55 years, and (D) aged 60 years. Notes: In this analytical sample, participants’ continuous health insurance enrollment was ascertained based on their healthcare utilization patterns (those who visited a medical institution were regarded as having health insurance enrollment for 180 days since the visit date).

**Table 2.** Percent change in regular dental visits after undergoing periodontal disease screening.

| **Analytic sample** | **DID estimates, % (95% CI\*)** |
| --- | --- |
| Total population (n = 4,270) | 12.1% (9.0–15.0) |
| Aged 20–35 years (n = 495) | 17.4% (8.0–27.1) |
| Aged 40–55 years (n = 2,750) | 11.5% (7.4–15.9) |
| Aged 60 years (n = 1,025) | 11.0% (4.7–16.9) |

Abbreviations: DID = difference-in-differences; CI = confidence interval.
Notes: In this analysis, participants' continuous health insurance enrollment was ascertained based on their healthcare utilization patterns (those who visited a medical institution were regarded to have the health insurance enrollment for 180 days since the visit date).
\* Obtained by wild cluster bootstrapping with 1,000 replications.

By age group, the increase in the prevalence of visiting dentists at least once in 180 days was sharper in those aged 20–35 years (from 56.6% in 1–180 days before to 84.8% in 1– 180 days after) than in those aged 40–55 years (from 61.3% in 1–180 days before to 81.3% in 1–180 days after) or 60 years (from 59.5% in 1–180 days before to 79.0% in 1–180 days after) (Figure 1). In the age group DID analysis, the corresponding percent changes were 17.4% (95% CI, 8.0–27.1) in those aged 20–35 years, 11.5% (95% CI, 7.4–15.9) in those aged 40–55 years, and 11.0% (95% CI, 4.7–16.9) in those aged 60 years (Table 2).

Figure 2 shows the prevalence of visiting dentists at least once every 180 days for 360 days after screening among those who had not visited dentists for 360 days before undergoing screening. In the total population, 69.6% of the participants visited dentists 1–180 days after screening. In addition, the prevalence was higher in the younger population (aged 20–35 years, 77.3%; aged 40–55 years, 73.1%; and 60 years, 58.2%). In all age groups, the prevalence at 181–360 days after screening declined from that at 1–180 days after screening (total population, 41.5%; age 20–35 years, 40.9%; age 40–55 years, 41.5%; age 60 years, 41.8%).

**Figure 2.**
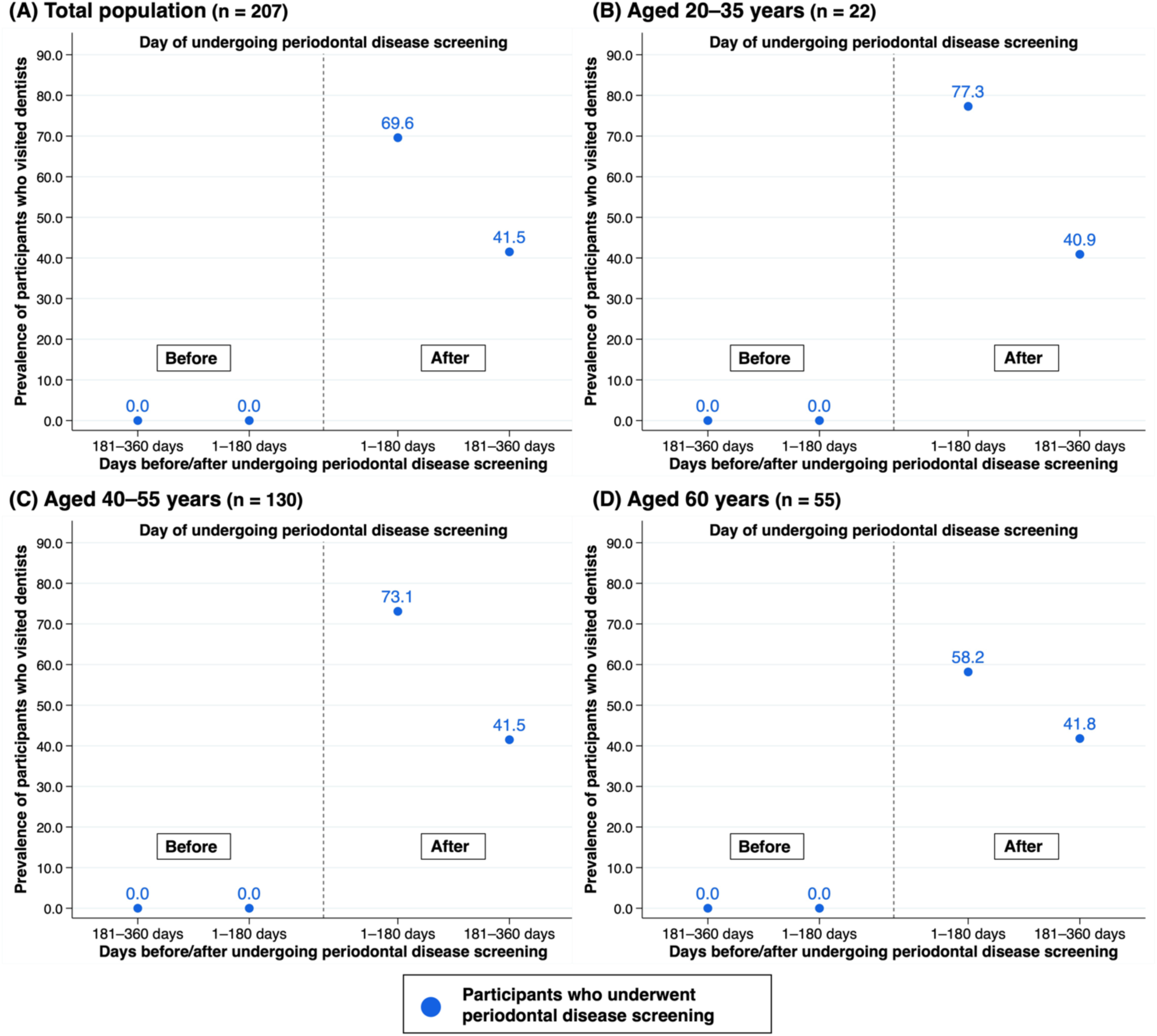
Prevalence of participants who visited dentists in 180 days during 360 days of undergoing periodontal disease screening in (A) total population, (B) aged 20–35 years, (C) 40–55 years, and (D) aged 60 years among those who had not visited dentists in 360 days before undergoing this screening. Notes: In this analytical sample, participants’ continuous health insurance enrollment was ascertained based on their healthcare utilization patterns (those who visited a medical institution were regarded as having health insurance enrollment for 180 days since the visit date).

Figure 3 shows the prevalence of each dental procedure that the participants who underwent screening received at least once in 1–180 or 181–360 days after the screening. In the total population, while the prevalence of patients who received cavity fillings declined from 1–180 days after screening (49.8%) to 181–360 days after screening (40.4%), the prevalence of patients who received dental calculus removal increased from 1–180 days after screening (69.3%) to 181–360 days after screening (71.7%) undergoing the screening. On the other hand, among the participants who had not visited a dentist 360 days before undergoing screening, the prevalence of those who received cavity fillings 1–180 days after screening (66.0%) was higher than that in the total population, but it was almost halved 181–360 days after screening (32.6%). In the secondary analysis, which ascertained the participants’ continuous health insurance enrollment using the health insurance register, the characteristics of the analytical samples were similar to those of the primary analysis (Supplemental Table 2). In addition, we observed similar trends in the prevalence of dental visits 180 days during 360 days before and after periodontal disease screening (Supplemental Figure 2). In the total population, the prevalence was 51.0% at 181–360 days before screening and 52.5% at 1–180 days before screening among those who underwent screening; it increased to 74.8% at 1–180 days and declined to 58.3% at 181–360 days after screening. In contrast, the prevalence in those who did not undergo screening was almost consistent throughout the study period (181–360 days before screening, 30.7%; 1–180 days before screening, 29.4%; 1–180 days after screening, 31.4%; 181–360 days after screening, 31.1%). In the analysis of those who had not visited dentists for 360 days before undergoing the screening, the prevalence at 1–180 days after screening was higher in the younger population (aged 40–55 years, 62.8%; aged 60 years, 54.9%) (Supplemental Figure 3).

**Figure 3.**
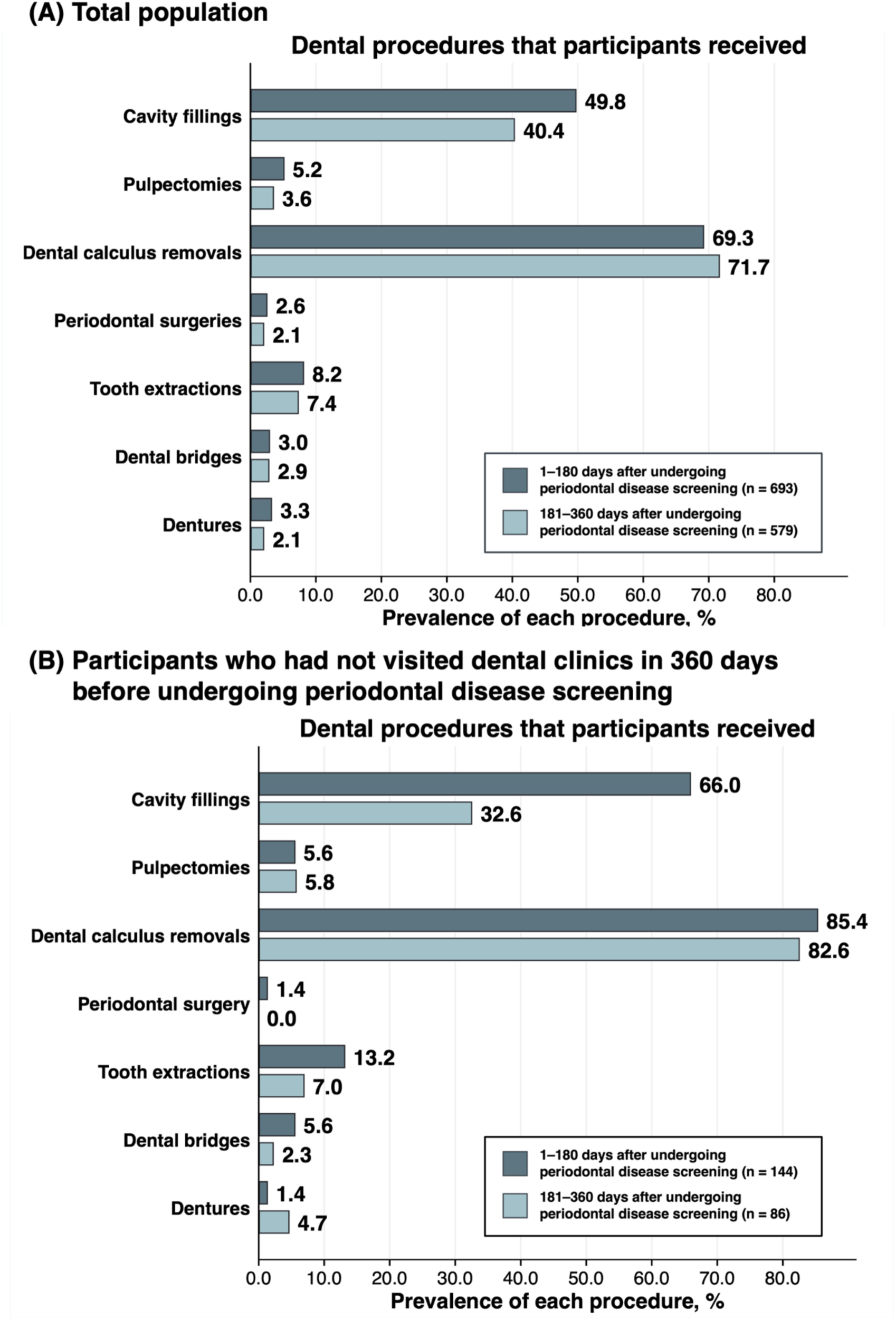
Prevalence of each dental procedure received in 1–180 days and 181–360 days after undergoing periodontal disease screening in (A) total population and (B) participants who had not visited dentists in 360 days before undergoing this screening.

## DISCUSSION

This study found that periodontal disease screening was associated with an increase in subsequent regular dental visits, including those who did not regularly visit dentists before screening. The increase in the frequency of regular dental visits after screening was more significant in the younger population.

Concerning the details of dental visits after undergoing the screening, this study revealed that nearly half of the participants had cavity fillings 1–180 days after the screening. The prevalence of those who received cavity fillings declined 181–360 days after screening. In contrast, the prevalence of dental calculus removal increased from 1–180 days to 181–360 days after screening. These results suggest that visits to the dentist for the treatment of caries noted during periodontal disease screening may have contributed to the increase in the prevalence of visits to the dentist on days 1–180 after screening. On the other hand, the prevalence of participants who underwent dental calculus removal was higher in 181–360 days than in 1–180 days after screening, but this cannot be simply interpreted as representing an increase in the prevalence of preventive dental visits. As the prevalence of visiting dentists declined from days 1–180 to days 181–360 after screening, it is possible that some people visited dentists for dental caries treatment and did not return after the treatment, which may have increased the prevalence of preventive dental visits. People who have experienced dental caries are known to be at a higher risk of developing it again^23^, so dental professionals should not only recommend dental visits during periodontal disease screening but also provide thorough oral hygiene education so that they can continue regular preventive dental visits after treatment.

In Japan, “universal dental health checks” are expected to be conducted in the next few years^10^. It is still unclear whether it will be an oral health screening program that provides annual individual screenings for the entire population, such as an expansion of periodontal disease screening, or an oral health campaign that promotes dental visits at least once a year.

If “universal dental health checks” would be an oral health screening program, because this study assessed the effectiveness of periodontal disease screening in late adolescent young older adults, we believe our findings may have important implications for developing the program. First, those who did not undergo periodontal disease screening were less likely to visit a dentist routinely than those who did. This suggests that, in developing “universal dental health checks”, a focused approach for those indifferent to their health (those in the pre-contemplation stage^24,25^) needs to be incorporated. Second, this study observed an increase in the prevalence of dental visits after screening, which declined over time. Based on the findings, “universal dental health checks” are expected to include a system that effectively promotes continuous dental visits. Further research is needed and incentives should be provided to those with continuous dental visits for preventive purposes.

This study has some limitations. First, given the low prevalence of people who underwent periodontal disease screening among the eligible people (around 5%)^26^, our study participants who underwent screening might have systemically higher health literacy than in those who did not. People with higher health literacy are more likely to follow recommendations from health professionals^27^; hence, it is possible that our study participants who underwent the screening were more likely to visit dentists regularly according to the advice received from the screening than those who did not. In this case, our estimates of the changes in the prevalence of visiting dentists may have been overestimated. Second, our study participants were restricted to the National Health Insurance enrollees, a public insurance scheme for those not employed. Enrollees included self-employed or part-time workers, retirees, and their dependents. Considering that full-time workers have to work for state-related reasons for not visiting dentists^28,29^, such as they cannot spare enough time on weekdays, caution is needed when generalizing our results to other populations^30^. Third, this study was based on municipal data. Although periodontal disease screening has been implemented in many municipalities and is carried out by the procedure set out^11^, there are some variations in its characteristics, such as eligible age, advertising method, and fees to undertake it. Hence, further studies using data from multiple municipalities are required to assess the generalizability of the results.

In conclusion, this study showed that screening for periodontal disease leads to subsequent regular dental visits among adults. In particular, younger individuals were more likely to visit a dentist after screening. We believe that our findings provide important insights for designing efficient oral health programs in the future.

## Appendix

Supplementary material can be found in the online version of the journal.

## Acknowledgments

We are grateful to the other investigators, staff, and participants of the LIFE Study for their valuable contributions. YT was supported by Nagoya University CIBoG WISE Program from MEXT.

## Author Contribution

YT, TK, MM, FM, HF, and KT conceived and designed the study. YT, MM, FM, and HF collected and checked data. YT analyzed the data. YT and KT wrote the first draft of the manuscript. KO, HF, and KT supervised and administrated the study. HF and KT acquired funding. All authors reviewed the manuscript, interpreted the data, and approved the final version of the manuscript. The corresponding author attests that all listed authors meet authorship criteria and that no others meeting the criteria have been omitted.

## Data Availability Statement

All data used in this study are not publicly available due to ethical or legal restrictions. For inquiries about the datasets used in this study, please contact the principal investigator of the LIFE Study, Dr. Haruhisa Fukuda, upon reasonable request. Codes used to generate the results in the manuscript may be available from the first author upon reasonable request.

