## Supplemental file for "Public oral health screening leads to regular dental visits: the LIFE Study"

**Title:**

**Supplemental Table 1.** Definition of Japanese original procedure codes for each procedure.

| | **Japanese original procedure codes** |
|---|---|
| Cavity fillings | 313000610, 313000710, 313000810, 313001210, 313001310, 313002010, 313002210 |
| Pulpectomies | 309002110, 309002210, 309002310, 309002410, 309002610, 309002710, 309002810, 309002910, 309011930, 309012030, 309017030, 309017130, 309012130, 309012230, 309017230, 309017330, 309012330, 309012430, 309012530, 309017430, 309017530, 309012730, 309012930, 309013130, 309013230, 309017830, 309013330, 309013430, 309013530 |
| Dental calculus removals | 304000410, 304000510, 304000610, 304000710, 304000810, 304000910 |
| Periodontal surgery | 310011610, 310011710, 310011810, 310011910, 310012010, 310012110, 310013010, 310013110, 310013210, 310013310 |
| Tooth extractions | 150110010, 150110110, 150110210, 150110310, 150110410, 310000110, 310000210, 310000310, 310000410, 310000510, 310000710 |
| Dental bridges | 313005010, 313005110 |
| Dentures | 313005310, 313005410, 313005510 |

**Supplemental Table 2.** Characteristics of analytic sample according to fiscal year when participants underwent periodontal disease screening.

| | **Overall** (n = 5,055) | | **Fiscal year 2017** | | | | **Fiscal year 2018** | | | |
|---|---|---|---|---|---|---|---|---|---|---|
| | | | **Participants who underwent periodontal disease screening** (n = 514) | | **Participants who did not undergo periodontal disease screening** (n = 2,056) | | **Participants who underwent periodontal disease screening** (n = 497) | | **Participants who did not undergo periodontal disease screening** (n = 1,988) | |
| | n | % | n | % | n | % | n | % | n | % |
| Sex | | | | | | | | | | |
| Male | 2,005 | 39.7 | 197 | 38.3 | 788 | 38.3 | 204 | 41.1 | 816 | 41.1 |
| Female | 3,050 | 60.3 | 317 | 61.7 | 1,268 | 61.7 | 293 | 59.0 | 1,172 | 59.0 |
| Age, years | | | | | | | | | | |
| 40 | 545 | 10.8 | 57 | 11.1 | 228 | 11.1 | 52 | 10.5 | 208 | 10.5 |
| 45 | 755 | 14.9 | 82 | 16.0 | 328 | 16.0 | 69 | 13.9 | 276 | 13.9 |
| 50 | 1,595 | 31.6 | 169 | 32.9 | 676 | 32.9 | 150 | 30.2 | 600 | 30.2 |
| 55 | 930 | 18.4 | 88 | 17.1 | 352 | 17.1 | 98 | 19.7 | 392 | 19.7 |
| 60 | 1,230 | 24.3 | 118 | 23.0 | 472 | 23.0 | 128 | 25.8 | 512 | 25.8 |
| Screening month | | | | | | | | | | |
| June | 197 | 19.5 | 103 | 20.0 | — | — | 94 | 18.9 | — | — |
| July | 183 | 18.1 | 91 | 17.7 | — | — | 92 | 18.5 | — | — |
| August | 125 | 12.4 | 62 | 12.1 | — | — | 63 | 12.7 | — | — |
| September | 110 | 10.9 | 63 | 12.3 | — | — | 47 | 9.5 | — | — |
| October | 144 | 14.2 | 60 | 11.7 | — | — | 84 | 16.9 | — | — |
| November | 252 | 24.9 | 135 | 26.3 | — | — | 117 | 23.5 | — | — |

Notes: In this analytic sample, participants' continuous health insurance enrollment was ascertained with the health insurance register.

**Supplemental Figure 1.** Prevalence of participants who visited dentists in 90 days during 360 days before and after undergoing periodontal disease screening in total population (n = 4,270).

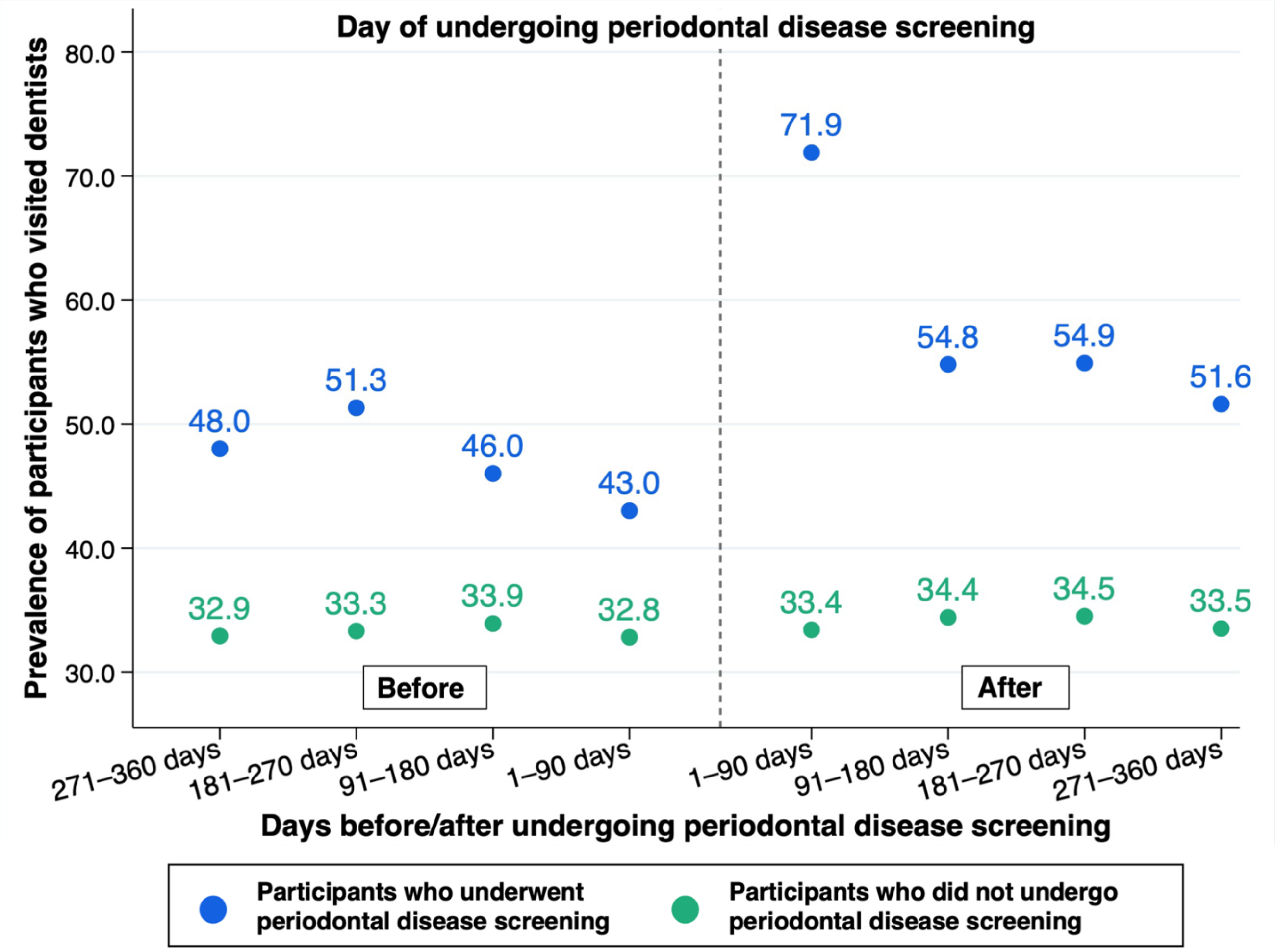

Notes: In this analytical sample, participants' continuous health insurance enrollment was ascertained based on their healthcare utilization patterns (those who visited a medical institution were regarded as having health insurance enrollment for 180 days since the visit date).

**Supplemental Figure 2.** Prevalence of participants who visited dentists in 180 days during 360 days of undergoing periodontal disease screening in (A) total population, (B) aged 40–55 years, and (C) 60 years.

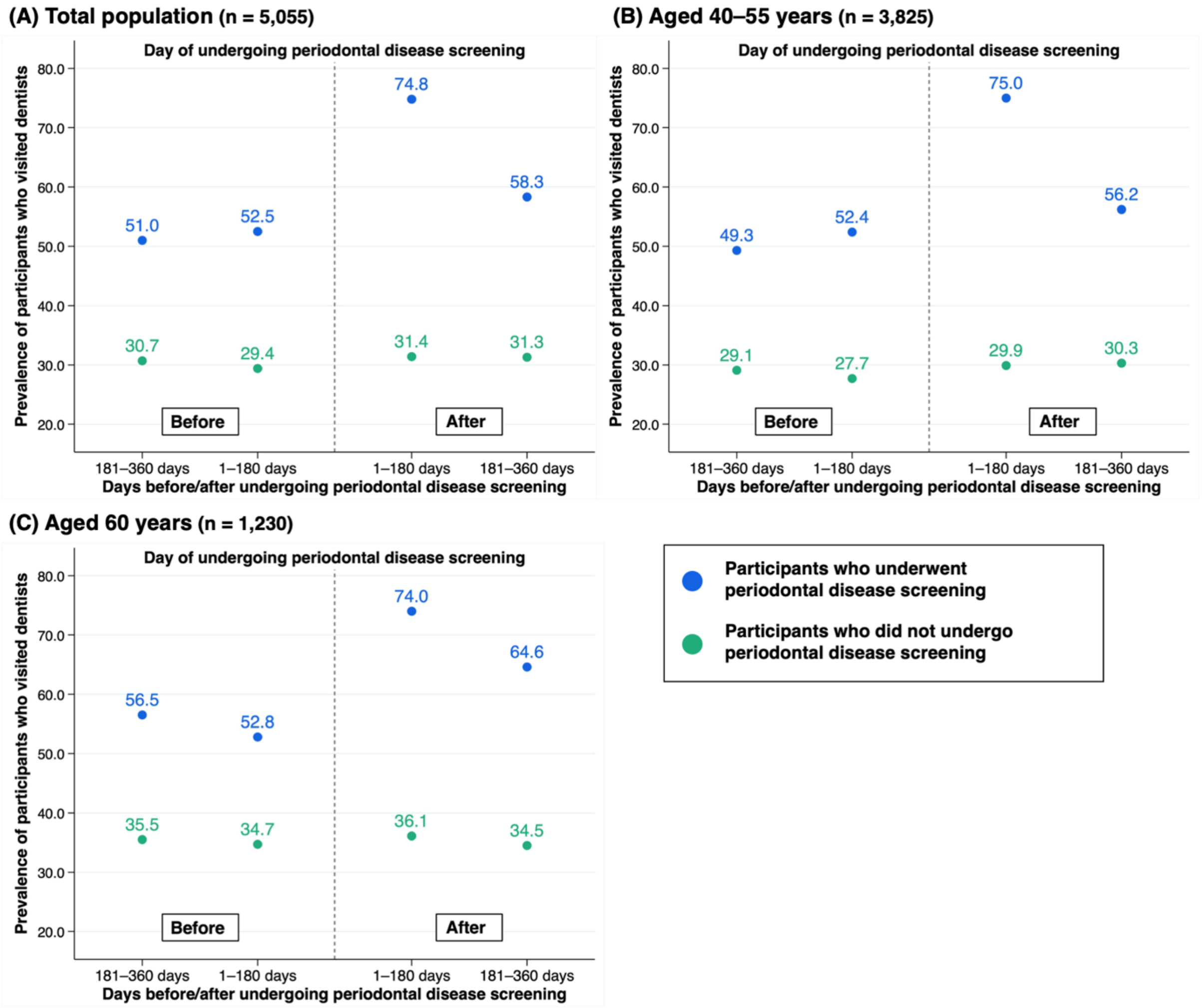

Notes: In this analytical sample, participants' continuous health insurance enrollment was ascertained with the health insurance register.

**Supplemental Figure 3.** Prevalence of participants who visited dentists in 180 days during 360 days of undergoing periodontal disease screening in (A) total population, (B) aged 40–55 years, and (C) aged 60 years among those who had not visited dentists in 360 days before undergoing this screening.

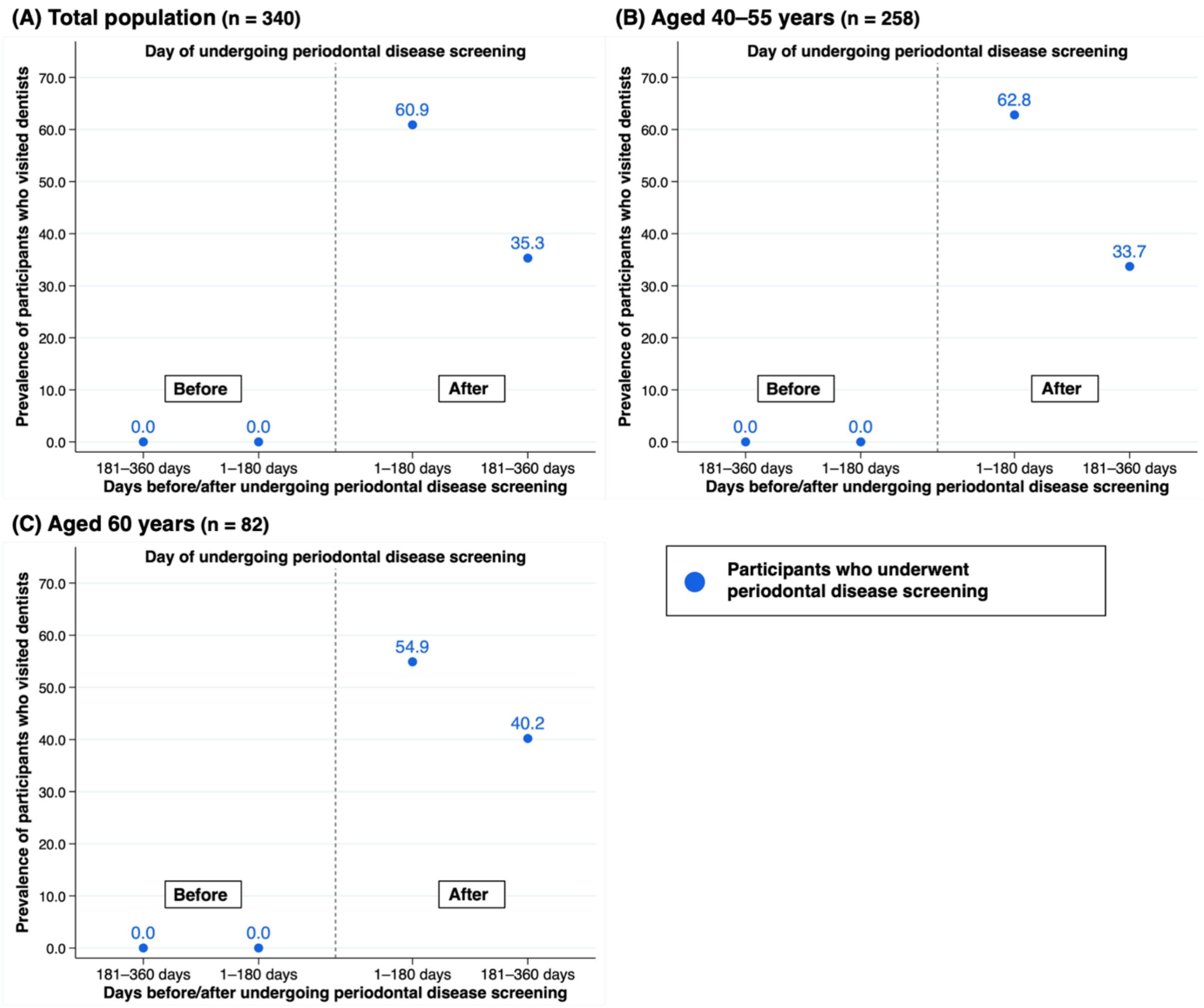

Notes: In this analytical sample, participants' continuous health insurance enrollment was ascertained with the health insurance register.
